# Effects of Human-Centric Lighting on Performance in the Attentional Network Test (ANT) and on Perceived Stress

**DOI:** 10.1101/2025.08.01.25332790

**Authors:** Nino Wessolowski, Tomasz Lukasz Ostrowski

## Abstract

The short-term effects of light on cognitive processes within the framework of Human Centric Lighting (HCL) remain underexplored and show inconsistent findings. In a laboratory study involving 789 participants, the influence of bright daylight white (BL), warm white (WL), and neutral light on executive functions was investigated using the Attention Network Test (ANT). BL emerged as the most beneficial condition overall, particularly enhancing the alerting component. In contrast, WL was repeatedly associated with negative effects on both alerting and executive attention. Unexpectedly, WL showed advantages in the orienting component, suggesting complex underlying mechanisms that are not yet fully understood. Future studies should examine lighting programs with greater contrast and incorporate physiological measures (e.g., pupillometry, EEG) as well as more differentiated psychological assessments. To explain these findings, the development of integrative models—such as the HCL-effect model proposed by Wessolowski—is essential. Light is more than a visual stimulus; it modulates attention and cognition. This insight should be systematically considered in the design of work, learning, and living environments.

## Introduction

The long-term effects of light on human physiology and behavior have been the focus of intensive interdisciplinary research. Since the discovery of the photosensitivity of retinal ganglion cells, the biological non-visual pathway of light processing has been well characterized [1]. Based on this foundation, light-based interventions can be designed to modulate circadian rhythms in a targeted manner and can be evaluated using validated methods [2].

In contrast, the short-term effects of light are theoretically less well understood and have yielded mixed findings in empirical research, particularly regarding attention and concentration [3]. The initial study by Wessolowski and Rahim [3] addressed this question and demonstrated a statistically significant small effect in favor of a brief bright-light intervention -lasting only a few minutes - on executive functions, as measured by the Attentional Network Test (ANT) [4] under daylight conditions.

Executive functions assessed by the ANT are associated with the activation of specific brain regions [4], which themselves are linked to circadian regulatory processes [5]. These findings therefore suggest that even short-term exposure to bright light may influence cognitive performance via the biological non-visual pathways - a mechanism previously well - documented primarily for long-term effects.

The pilot study by Wessolowski and Rahim [3] contributed to narrowing this research gap but, due to its limited sample size, did not meet the methodological standards of a fully powered laboratory experiment. Consequently, the study was extended in subsequent years. The present article reports the results of this expanded investigation.

### 1.1 Pathways of the effect of light on human behavior

The effects of light on human behavior - particularly on attention and concentration - can be explained by three central theoretical pathways or hypotheses [3,6,7]:

1. **The visual pathway** refers to the enhancement of visual perception through increased illuminance [8]. In the present study, this pathway plays a subordinate role, as screen brightness was held constant and only ambient lighting was manipulated. While ambient lighting can influence the perception of screen content, such effects are likely to be minimal under the given experimental conditions. In studies involving paper-based attention tests, however, the visual pathway may have had greater relevance.
2. **The neurobiological non-visual pathway** is based on the effects of light on intrinsically photosensitive retinal ganglion cells (ipRGCs), which are connected via the retinohypothalamic tract to the suprachiasmatic nucleus—the master pacemaker of the circadian system [1,9,10]. This pathway influences physiological arousal and activity levels through hormonal regulation, particularly the suppression of melatonin secretion and modulation of cortisol levels [5]. Although the temporal dynamics between light exposure, neuroendocrine responses, and observable behavior are not yet fully understood [11,12], recent findings indicate that even brief light exposure during the day can activate brain regions associated with the circadian system [13]. Moreover, EEG studies suggest that light can modulate cortical alpha activity—a neural marker associated with vigilance and attentional processes [14,15].
3. **The psychological pathway** includes mood-related and conditioned responses to light. Bright, daylight-like lighting is associated in many cultures with activity, alertness, and positivity and may thus promote both subjective well-being and cognitive performance [6,11,12]. In contrast, dim lighting is often associated with passivity or negative emotions such as anxiety [16]. Studies have shown that certain light colors or color temperatures perceived as pleasant are linked to improved performance in tasks requiring selective attention and problem solving [11]. Interindividual differences may also result from conditioning or different learning experiences [3,6,7].

The effects of light unfold in interaction with individual factors (e.g., age, sex, ocular anatomy [11,12,17]) and situational contexts (e.g., spatial environment, social setting [18]). In particular, individual arousal levels appear to play a central mediating role in how light influences behavior and experience [19,20]. Against this background, Wessolowski [6] proposes an integrated model of light effects that accounts for these moderating influences. The model conceptualizes light effects initially as physiological responses, which subsequently lead to changes in behavior and experience (see Fig 1).

**Fig 1.**
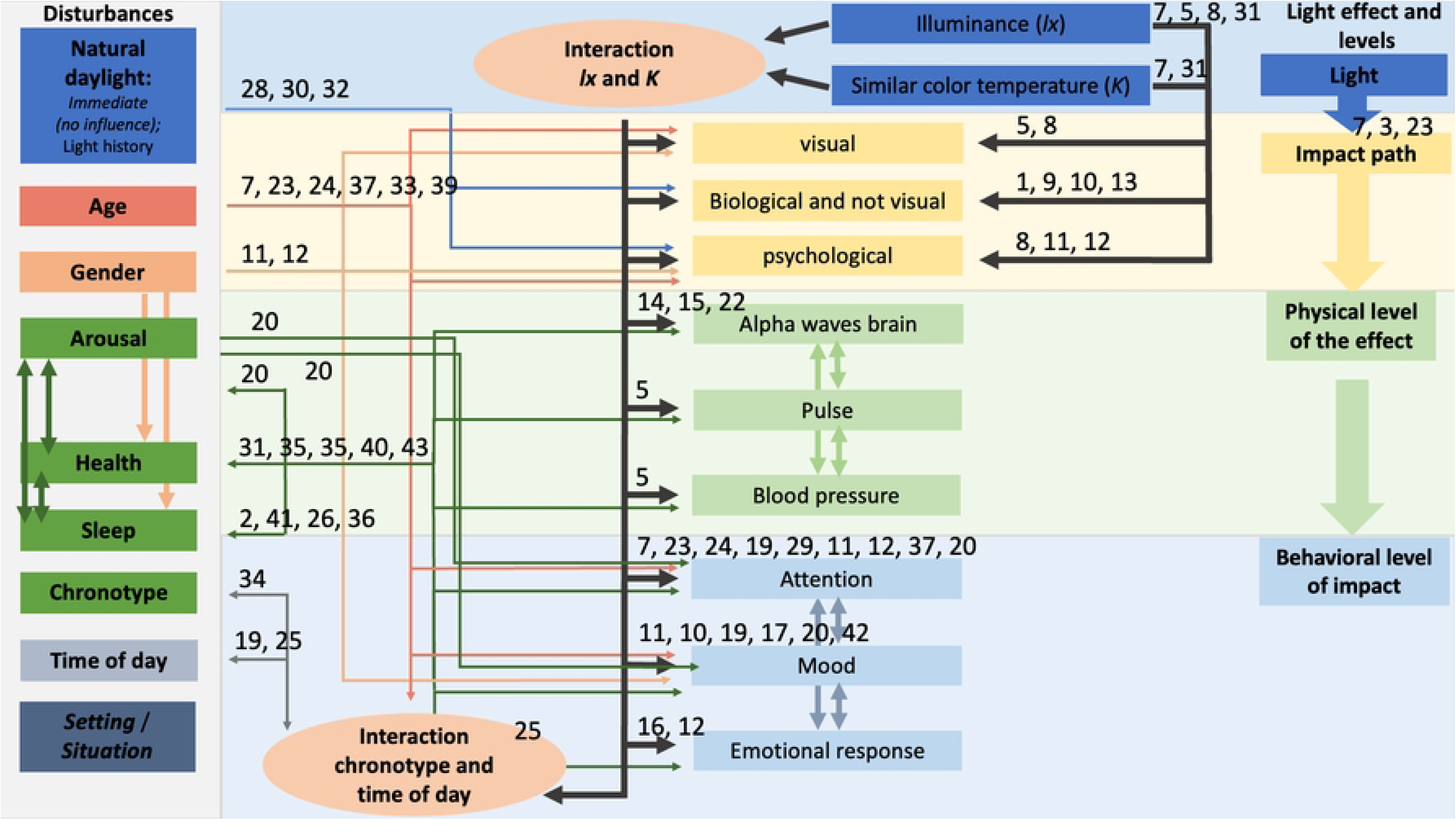
Theoretical base model by Wessolowski [6], which can be empirically tested, optimized using structural equation modeling, and conceptually expanded. The numbers shown in the figure refer to sources listed in the reference section.

Despite numerous empirical findings, the relationship between light as a circadian zeitgeber and short-term behavioral changes throughout the day remains insufficiently understood. The aim of the present study is therefore to provide further empirical evidence for the immediate effects of daylight exposure on behavior and experience—specifically, attention and concentration.

### 1.2 Previous studies on the attentional effects of light

In a meta-analysis by Golmohammadi et al. [45], 19 studies on the effects of light on attention were synthesized. The results indicate that light sources with shorter wavelengths, higher intensity, and higher color temperature suppress melatonin production, increase alertness, reduce fatigue, enhance attention, and improve reaction time. According to this meta-analysis, bright daylight in the morning - characterized by short wavelengths, high intensity, and prolonged exposure - is the most effective type of light for regulating psychological, biological, and cognitive processes.

The findings demonstrate that light is a strong modulator of non-visual cognitive performance. Wavelength, color temperature, and light intensity influence neural responses to cognitive processes such as attention and reaction speed. The authors recommend that these parameters - alongside individual and environmental factors - should be considered when designing and implementing light interventions.

Another meta-analysis by Mu et al. [46], which included 29 studies examining the effects of light on alertness, also supports the beneficial effects of light interventions on both subjective and objective measures of alertness in healthy individuals. Cold light was found to be more effective than warm light in improving both parameters. Furthermore, light exposure increased subjective alertness during both daytime and nighttime.

Overall, the meta-analyses point to a positive effect of bright daylight, especially light with a high blue component. However, the reviewed studies primarily investigated longer exposure durations - typically more than 20 minutes [46] or even several hours [45] - and thus significantly exceeded the exposure duration used in the present study. It should also be noted that some of the included studies reported increased stress levels among participants, while others did not [46]. Moreover, the evidence base regarding attention and concentration is less consistent than these meta-analyses might suggest at first glance. Individual studies report divergent effects of light on various subcomponents of attention, as illustrated below through our own research and thematically related work from other groups.

Previous studies investigating the specific effects of light on attention generally present a differentiated, predominantly positive picture - particularly in children and adolescents. A field study by Barkmann et al. [24] examined the effects of artificial daylight (1060 lx, 6500 K) compared to standard lighting (300 lx, 4000 K) in primary and secondary schools (n = 110). Under daylight conditions, the number of errors in the d2 test was significantly reduced (small effect size), and reading performance improved significantly (medium effect size). No significant effects were found for processing speed or concentration performance (CP).

A complementary laboratory study by Wessolowski [7] involving 95 university students also found significant effects (small effect size), including a reduction in errors (especially omissions) and, in contrast to the field study, an improvement in processing speed.

A comprehensive study by Sleegers et al. [37] comprised three successive experiments involving approximately 190 primary school children. In the first two field studies, bright light conditions (1000 lx, 6500 K) led to a significant improvement in concentration performance (CP) and a reduction in d2 errors compared to standard lighting (300 lx, 3000–4000 K). However, the third experiment - a randomized laboratory study - did not yield significant effects, possibly due to the artificial setting and short exposure duration.

In the study by Auras et al. [23] an A-B-A-B design was applied in a child and adolescent psychiatric clinic with 30 patients (mean age = 11.5 years). Under the daylight condition (5312 K, 793 lx), significant improvements were observed in concentration performance and processing speed, but not in error rate. Interestingly, participants subjectively reported poorer concentration, as well as increased stress and fatigue.

Finally, Weitbrecht et al. [47] investigated the effects of different light color temperatures (3000 K, 4500 K, 6000 K) under constant illuminance (1000 lx) in a sample of 50 students and professionals. No significant differences were found in the d2-bq test. However, warm light (3000 K) was associated with higher creativity, while higher-blue-content light (4500 K, 6000 K) showed a trend toward improved attention, particularly in a word-based attention test.

In summary, most studies report significantly positive effects of bright daylight on concentration performance. However, these effects vary depending on study design, setting, and measurement tools. The relationship between light interventions and subjective stress remains unclear, as do the underlying mechanisms of short-term light effects.

To address these discrepancies in the literature, explore the influence of subjective stress perception, gain a deeper understanding of the underlying mechanisms, and examine the transferability of effects to adults, Wessolowski and Rahim [3] conducted a study on the short-term effects of light on selective attention. A total of 95 participants completed the Attention Network Test (ANT) under conditions of bright daylight versus warm white light. The study focused on executive attention, as this construct best corresponds to the selective attention aspect measured by the ANT short form. Bright daylight significantly improved executive attention (small effect size), indicating that bright daylight can acutely enhance executive functioning and, consequently, selective attention in young adults.

However, this pilot study did not reach the sample size of n = 788 required for sufficient statistical power. Therefore, the present study was conducted to extend this line of research.

### 1.4 Questions and hypotheses of the present study

The present study aims to investigate the effects of light on young adults in a more differentiated manner. In particular, it seeks to clarify how light influences executive attention. To this end, executive attention was assessed using the short form of the Attention Network Test (ANT) [4], allowing for a more process-oriented analysis compared to previous studies that employed the d2 test to measure attention [48]. The d2 test captures attention performance in a more global and less process-specific way, which may partly explain the inconsistent findings reported in earlier research. We assume that the congruency score (executive attention) in the ANT corresponds most closely to the construct of selective attention as operationalized in the d2 test.

In summary, this study addresses three central research questions:

1. Does bright daylight enhance executive attention performance compared to neutral and dimmed warm light conditions?
2. What effects can be observed in the alerting and orienting components of attention?
3. Do the three lighting conditions differ in their effects on subjective stress perception?

## Materials and Methods

The methodological design of the present study was largely adopted from the pilot study described in Wessolowski and Rahim [3], with only minor modifications - namely, the introduction of a neutral lighting condition. For the sake of completeness and to enhance comprehensibility, the methodological framework is summarized below.

### 2.1 Sample

The required sample size was determined through an a priori power analysis using G*Power [49]. Assuming small effect sizes and a significance level of p < .05, the analysis indicated an optimal sample size of n = 788. Due to the logistical complexity of the laboratory setup, collecting a sample of this size required data collection over several years. Initial findings based on data from the first semester (n = 91; April 20, 2021 – May 16, 2021) were published as a pilot study by Wessolowski and Rahim [3]. Data collection continued until December 12, 2023 (semester 6), and the full dataset is analyzed and reported in the present study.

Participants were recruited from a pool of Bachelor’s and Master’s students, as well as their families and social networks. A total of *n* = 789 individuals participated and were included in all analyses, as no missing data were recorded. The sample consisted predominantly of psychology students (67.6%), and a majority of participants were female (65.4%) (see Table 1).

**Table 1.** Sample information.

|  | BL | NL | WL | total | Significance between groups |
| --- | --- | --- | --- | --- | --- |
| n (%) | 335<br>(42.6) | 142<br>(18.0) | 312<br>(39.5) | 789<br>(100.0) | $X^2 = 84.510$, df = 2, p = .000* |
| age in years M (SD) | 24.7<br>(6.65) | 24.1<br>(3.89) | 24.7<br>(6.63) | 24.6<br>(6.24) | U= 3.092, df = 2, p = .213 |
| sex f/m in % | 66.9/33.1 | 57.6/42.5 | 67.3/32.7 | 65.4/34.6 | $X^2 = 4.601$, df = 2, p = .100 |
| Students of psychology in % | 70.7 | 58.5 | 68.3 | 67.6 | $X^2 = 6.999$, df = 2, p = .030* |

Notes. BL = bright daylight, NL = neutral-white light, WL = dim warm light, U = Mann-Whitney-U-Test (two tailed), $X^2$ = Chi-Square Distribution.

As previously observed by Wessolowski and Rahim [3], psychology students were not evenly distributed across the experimental groups despite random assignment; the reason for this imbalance remains unknown. In addition, due to violations of the assumption of normality, no t-tests were conducted to compare the groups. Instead, Mann–Whitney U tests or Chi-Square tests were used depending on the level of measurement (see Table 1). The unequal group sizes across lighting conditions can be attributed to the fact that the neutral lighting condition was introduced only in the fourth semester (January 2023).

None of the control variables were significantly correlated with the ANT outcome measures (i.e., the dependent variables). Therefore, they were not included as covariates in the final analyses.

### 2.2 Design and Materials

The study was conducted in the laboratory of the Medical School Hamburg (MSH) using four identical, sound-insulated, and ventilated test cabins (dimensions: 180 cm in length, 220 cm in height) with light grey interior walls. Each cabin contained a 72 × 35 cm window located behind the participant, to the left of the entrance door. These windows faced away from the external laboratory windows (north-east orientation), allowing minimal natural daylight to enter the cabins (maximum recorded across all measurements: 30 lx; see control variables). Each cabin was furnished with two chairs, a table, a computer, a screen, and two lamps (see Figs. 2 - 4).

**Fig. 2:**
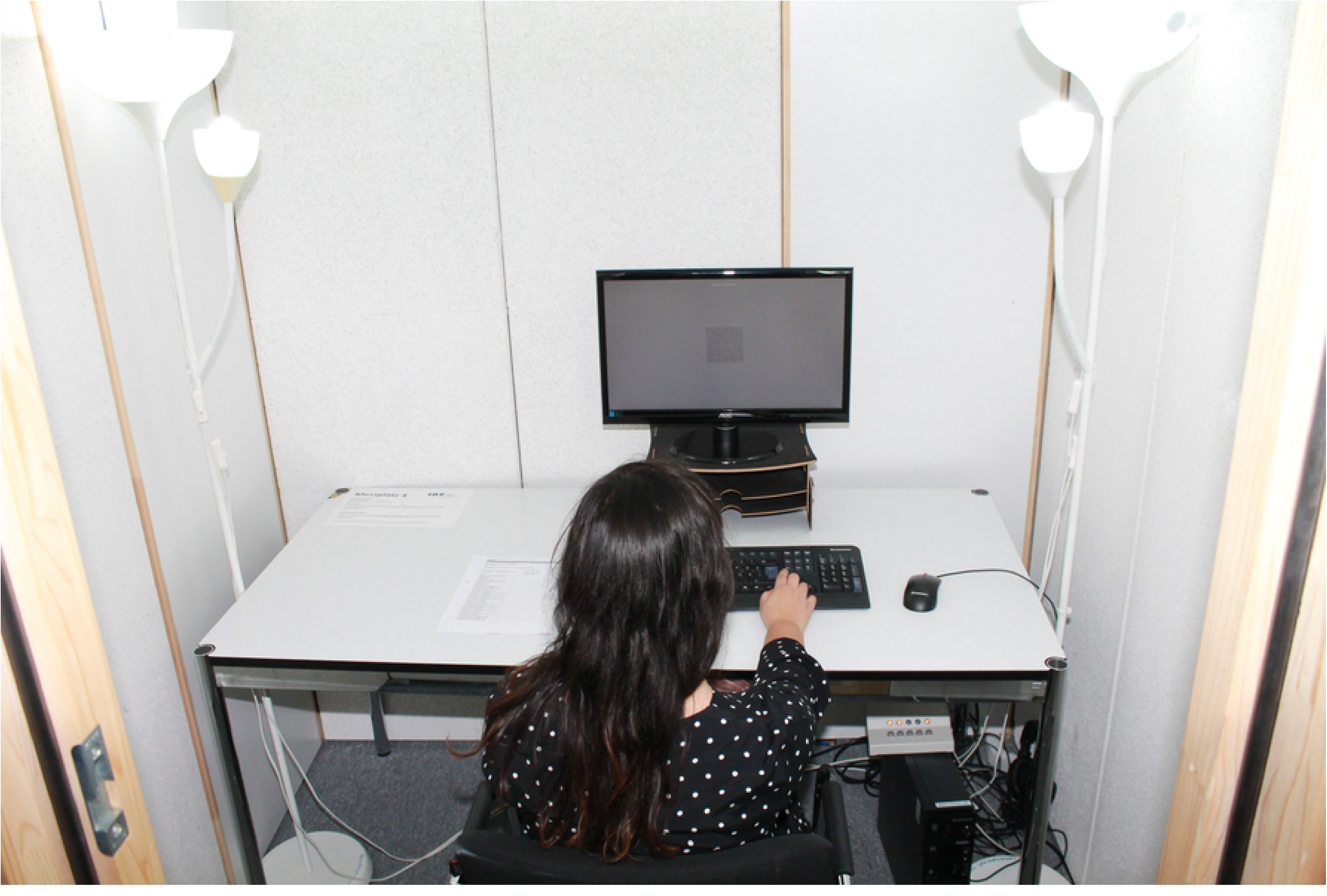
Laboratory cabin under lighting condition: bright daylight (BL).

**Fig. 3:**
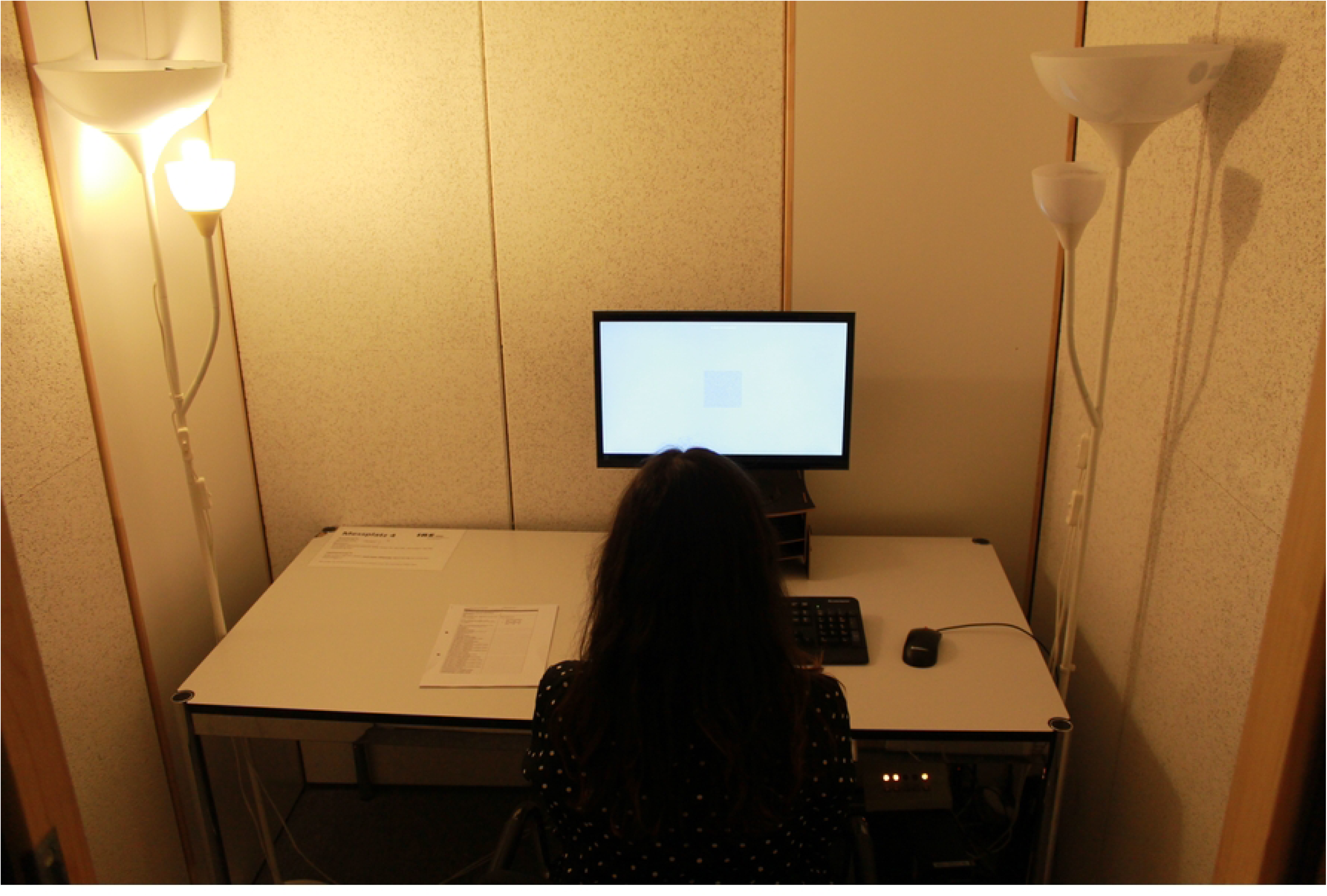
Laboratory cabin under lighting condition: dim warm light (WL).

**Fig. 4:**
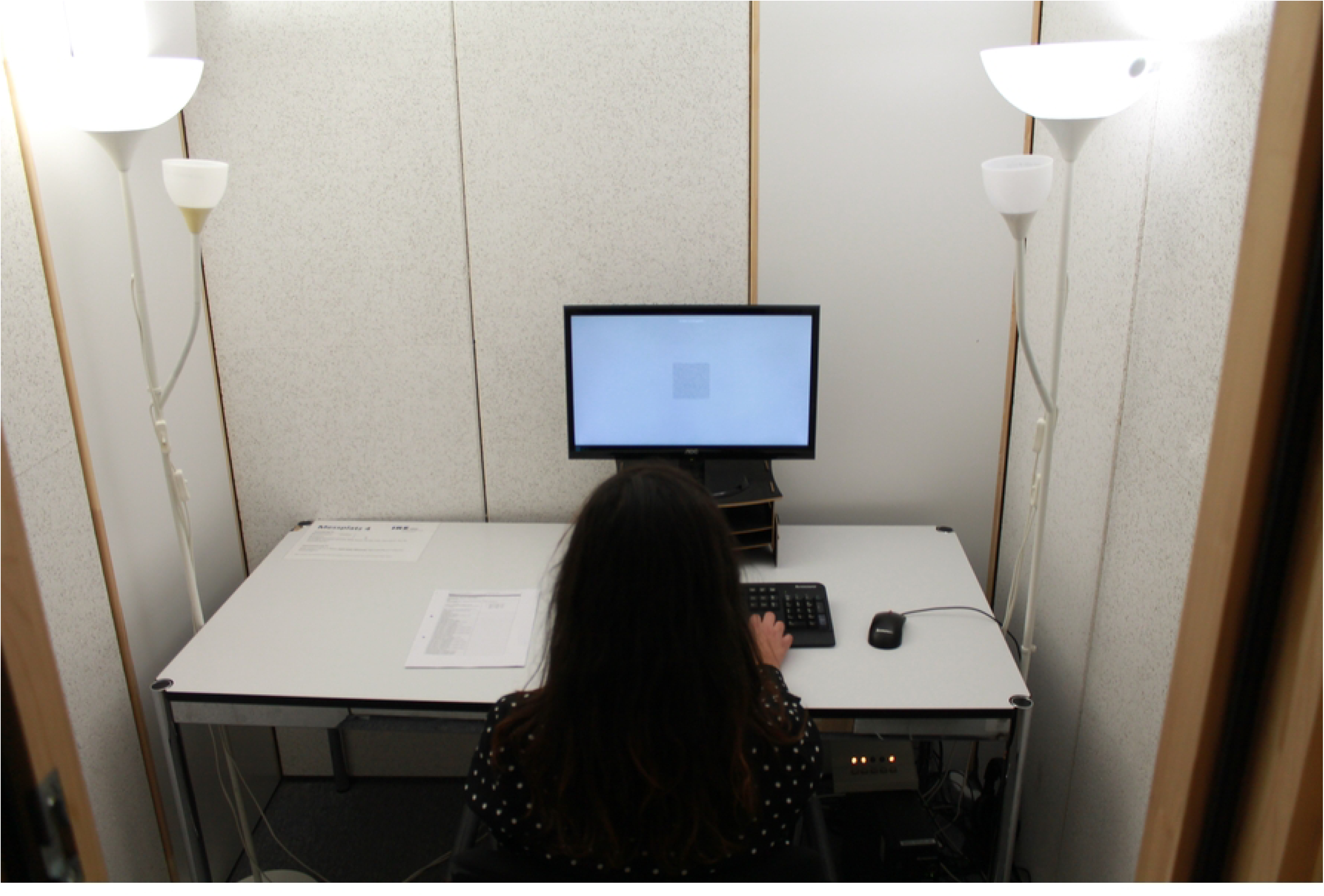
Laboratory cabin under control lighting condition: neutral light (NL).

The technical setup consisted of a Lenovo PC (Intel® Pentium® CPU G3220 @ 3.00 GHz, 4.0 GB RAM), running Windows 7 Professional (64-bit), connected to a 22” LCD monitor. The Attention Network Test (ANT; see below) was administered via the experimental software E-Prime 2.0 (Psychology Software Tools, Inc.).

As shown in Figure 2, the bright daylight (BL) condition was realized using two standard floor lamps (Ikea Not) equipped with compact fluorescent bulbs (Philips T65 Softone Cool Daylight E27 20W 865 and Philips Genie Cool Daylight E14 8W 865), along with a surface-mounted light fixture (Luxero T5LL-21 W) containing a fluorescent tube (FSL 21W 865/0).

In contrast, the dim warm-white (WL) condition (Fig. 3) used the same floor lamp fixtures but with a different bulb: a Philips Tornado E14 12W 827 compact fluorescent lamp. In both lighting conditions, the adjustable lamp arms (E14 sockets) were fixed to the interior cabin wall, with the light beam directed toward the ceiling to ensure diffuse, indirect illumination. In the third lighting condition neutral light (NL), both uplighters were equipped with a Taloya A60 E27 9W 840 bulb each (Fig. 4). To quantify lighting parameters, each cabin was equipped with an ALP-01 measuring device from Asensetek (Lighting Passport). Illuminance and correlated color temperature (CCT) were measured horizontally at the work surface. Further details on the operationalization of lighting conditions and the photometric measurements are discussed in 4.2 (*Limitations*).

Under the bright daylight (BL) condition, average values were 517.0 lx and 5417 K- indicating high light intensity and a daylight-like color temperature with a blue component (see Fig. 2). Under the dim warm-white (WL) condition, the average was 67.5 lx and 2628 K - producing a low-light environment with a warm-white color component (see Fig. 3). In the “neutral light” (NL) condition, the average values were 160.8 lx and 4133 K. The overall impression of the cabin under this lighting is shown in Figure 4.

The contribution of the monitor to overall cabin lighting was minimal, with an average illuminance of 14.6 lx and a color temperature of 6466 K.

### 2.2 Variables

In this study, the ANT-Short was used [4]. Participants were instructed to respond as quickly as possible to the direction of a central arrow displayed on a computer screen by pressing the left or right mouse button with their corresponding thumb. The **Conflict Effect** (executive attention) was calculated as the difference in reaction times between trials with congruent and incongruent flankers [4]. The corresponding stimuli are illustrated in Figure 5.

**Figure 5.**
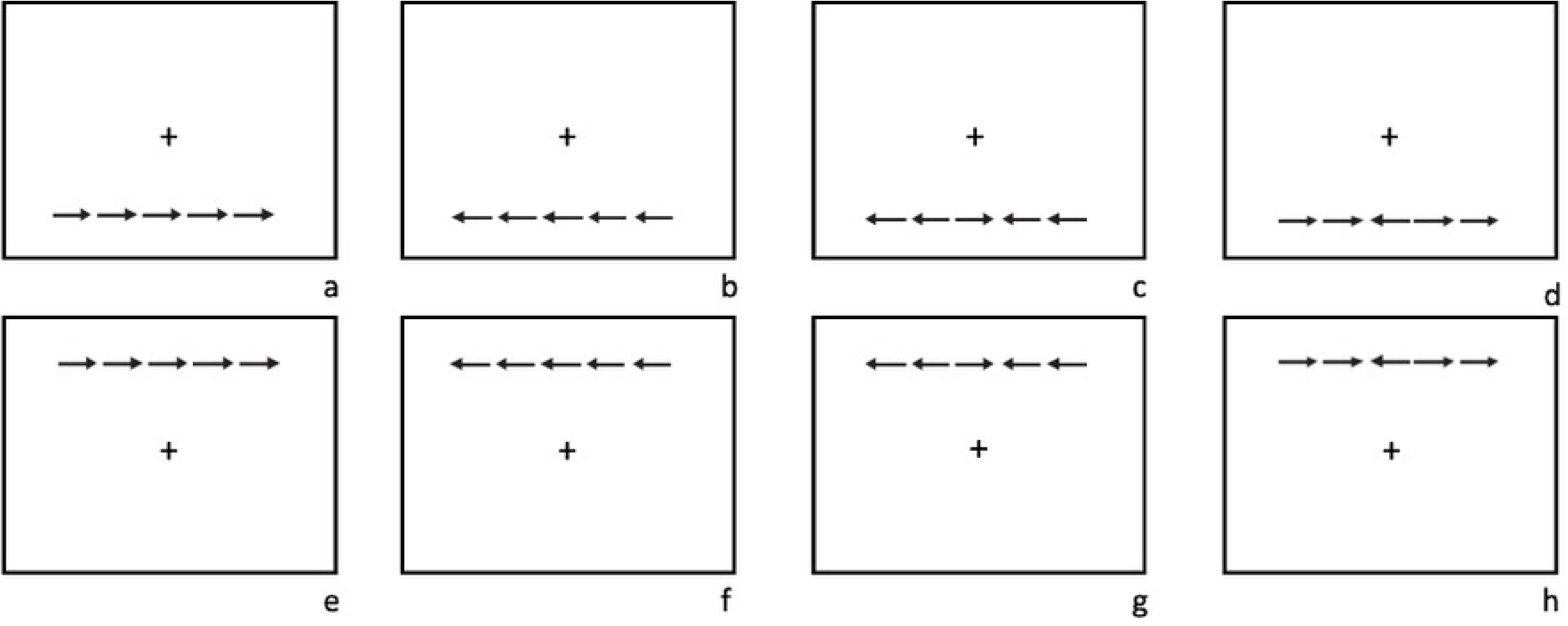
(a–h): Congruent bottom (a–b), incongruent bottom (c–d), congruent top (e–f), incongruent top (g–h).

The **Orienting Effect** was calculated as the difference in reaction times between trials with a center cue and a spatial cue [4]. These stimuli are illustrated in Figure 6.

**Figure 6.**
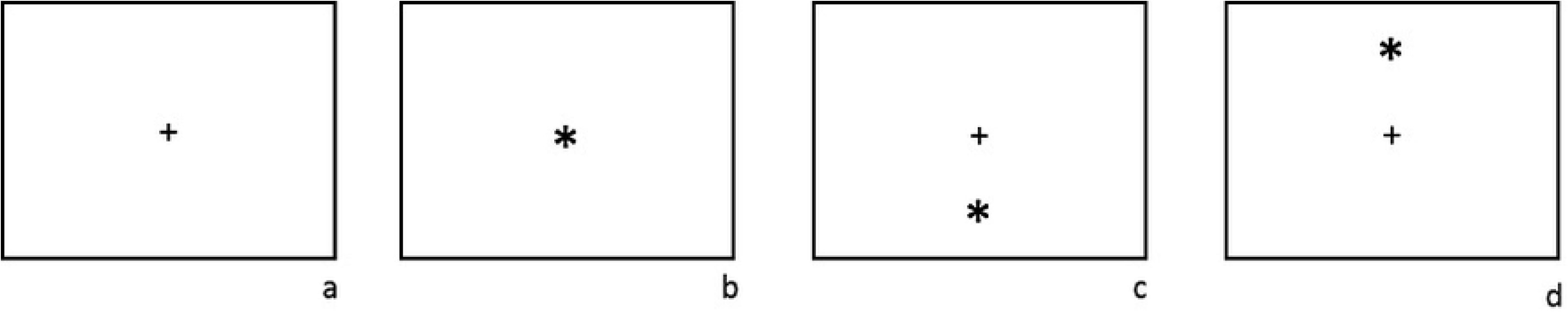
(a–d): No cue – fixation cross only (a), center cue (b), spatial cue bottom (c), spatial cue top (d).

The **Alerting Effect** was calculated as the difference in reaction times between trials with no cue and a center cue [4]. These stimuli are also illustrated in Figure 6.

Since lighting conditions might influence each condition of the ANT in the same way, all conditions (congruent, incongruent, informative/spatial, center, no cue) were analyzed individually, even though this is not explicitly prescribed in the ANT manual.

In line with previous findings, ratings of acute stress were also obtained. The **Rest vs. Restlessness** subscale of the Multidimensional Mood State Questionnaire (MDBF) [50] was used for this purpose.

Additionally, several control variables were assessed using short, self-formulated scales: long-term stress, caffeine intake, chronotype, previous night’s sleep duration, nationality, native language, use of glasses/contact lenses, handedness (left/right), general health status, and presence of chronic diseases.

Further contextual variables such as sunshine duration, day length [51], room temperature, date, and time of day were recorded. Moreover, pulse and blood pressure were measured using digital monitors (Sanitas SBC 15).

### 2.4 Procedure

Data collection took place between April 20, 2021, and December 12, 2023. The allocation of light conditions was randomized by coin toss. Starting from semster 4 (until January 2023), an initial coin toss determined whether participants would receive the control or intervention lighting. If assigned to the intervention group, a second coin toss was used to decide which of the two intervention conditions would be applied. Despite this procedure, the control group remained underrepresented overall (see Table 1). Accordingly, the luminaires were therefore fitted with the appropriate lamps for lighting condition, switched on and the room temperature measured. This procedure was determined in order to create comparable initial conditions and to allow the light effect to work. The adult participants were informed in both written and verbal form about the study and gave their written consent. The participants were first asked to complete questionnaires. This should result in a total acclimatization and exposure of about 10 minutes before the actual experiment. The experimenter stayed in the cabin during the instruction of the ANT and the test trial. Afterwards pulse and blood pressure were measured. Then the experimenter left the cabin and the participants completed the experiment, consisting of three ANT blocks each containing 48 trials. Each of these blocks was followed by a response on the scale of calm / restlessness. Pulse and blood pressure values were measured for the second time and afterwards the participant was send-off.

### 2.5 Ethics

The experiment was conducted in accordance with the Declaration of Helsinki and was approved by the Ethics Committee of Medical School Hamburg (**MSH- 2020105)**.

### 2.6 Statistics

Analysis of variance (ANOVA) procedures were originally planned to test the hypotheses. The assumptions for these analyses had been successfully evaluated in the pilot [3] and the planned procedures were deemed appropriate. However, assessment of the full dataset in this study revealed that the assumption of normality could not be confirmed, as determined by the Smirnov test, the Kruskal-Wallis test, and a less stringent evaluation based on skewness and kurtosis [52]. Log transformation using the natural logarithm also failed to normalize the data. Therefore, the ANT scale data were analyzed using the Mann-Whitney U test, which does not assume normality, conducted with SPSS 27 on macOS 14.6.1. In contrast, the subjective stress ratings showed a normal distribution within each light intervention condition, allowing for the planned ANOVA procedures to be carried out using SPSS. Using two-side testing and a local error probability of 5%, at least small effects were anticipated. A Huynh-Feldt correction was used when the sphericity assumption was violated.

## Results

### 3.1 Analysis of the ANT

Table 2 presents the differences in reaction times (in milliseconds) across the three lighting conditions for the scales of the Attentional Network Test (ANT).

**Table 2.**
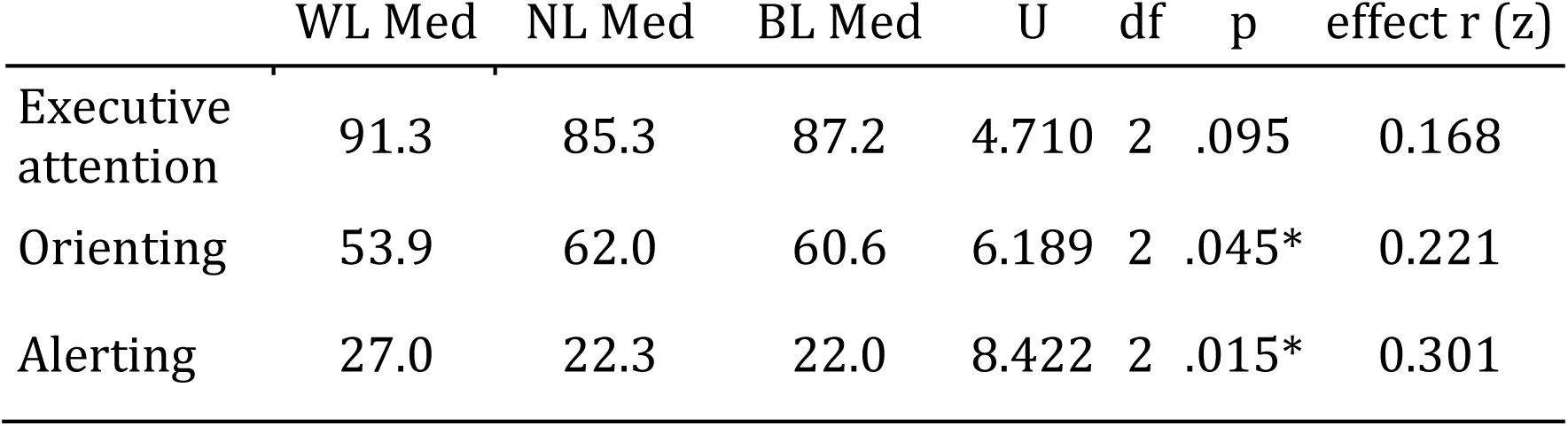
Analysis of the ANT (reaction time difference in milliseconds, n = 781).

For the **executive attention** component, a small and non-significant effect was observed, χ²(2) = 4.71, p = .095, r = .168. Reaction times under neutral light (NL, Mdn = 85.3 ms) and bright daylight white light (BL, Mdn = 87.2 ms) were similar and tended to differ from the warm white light condition (WL, Mdn = 91.3 ms), with NL showing a slight advantage. On average, participants achieved the poorest scores in executive attention tasks under WL. Compared to WL, NL resulted in a 6.0% improvement, and BL in a 4.1% improvement.

For the **alerting component**, a significant medium-sized effect was found, χ²(2) = 8.42, p = .015, r = .301. The shortest reaction times were recorded under bright daylight white light (BL, Mdn = 22.0 ms), followed by neutral light (NL, Mdn = 22.3 ms), and warm white light (WL, Mdn = 27.0 ms). WL consistently produced the poorest (reference) values. Compared to WL, reaction times improved by 5.0% under BL and by 4.7% under NL.

For the **orienting component**, a small but significant effect was observed, χ²(2) = 6.19, p = .045, r = .221. In this case, the shortest reaction times occurred under warm white light (WL, Mdn = 53.9 ms), followed by bright daylight white (BL, Mdn = 60.6 ms), and neutral light (NL, Mdn = 62.0 ms). NL showed the poorest values, while BL was 1.4% faster and WL showed an 8.1% improvement.

In addition to the ANT scale scores, which are based on differences between demanding and optimal conditions, the individual trial conditions were also analyzed separately. No significant effects were found in these analyses (see Table 3). The assumptions of normal distribution and homogeneity of variance required for the analysis of variance procedures had been successfully tested in advance.

**Table 3.**
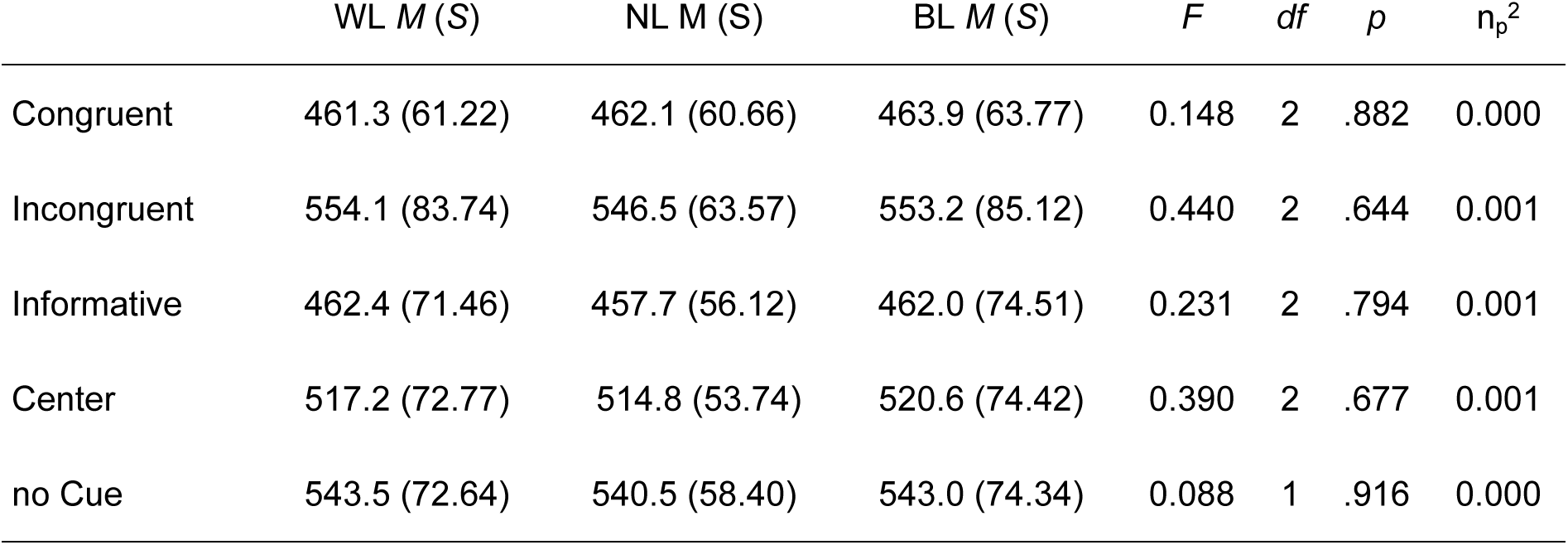
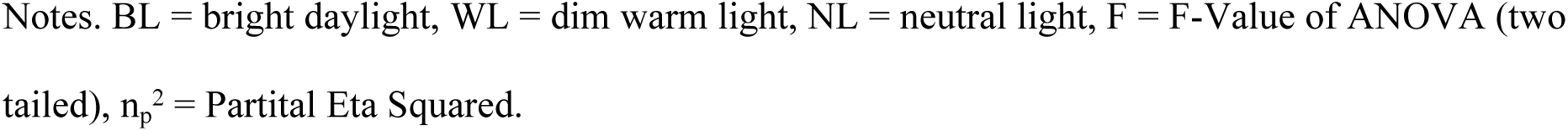
Analysis of the ANT conditions (reaction time in milliseconds, n=781).

| | WL M (S) | NL M (S) | BL M (S) | F | df | p | $\eta_p^2$ |
| --- | --- | --- | --- | --- | --- | --- | --- |
| Congruent | 461.3 (61.22) | 462.1 (60.66) | 463.9 (63.77) | 0.148 | 2 | .882 | 0.000 |
| Incongruent | 554.1 (83.74) | 546.5 (63.57) | 553.2 (85.12) | 0.440 | 2 | .644 | 0.001 |
| Informative | 462.4 (71.46) | 457.7 (56.12) | 462.0 (74.51) | 0.231 | 2 | .794 | 0.001 |
| Center | 517.2 (72.77) | 514.8 (53.74) | 520.6 (74.42) | 0.390 | 2 | .677 | 0.001 |
| no Cue | 543.5 (72.64) | 540.5 (58.40) | 543.0 (74.34) | 0.088 | 1 | .916 | 0.000 |

Notes. BL = bright daylight, WL = dim warm light, NL = neutral light, F = F-Value of ANOVA (two tailed), $\eta_p^2$ = Partital Eta Squared.

### 3.2 Stress

To assess subjective stress perception, values from the MBDF subscale Calmness/Restlessness were tested for normal distribution and subsequently analyzed using repeated-measures ANOVA. No significant or meaningful main effect of time was found, F(6) = 2.08, p = .104, η²= .003. There was also no significant effect of group, F(2) = 1.87, p = .156, η² = .005 (not further relevant to the present study), nor a significant or meaningful interaction between the critical factor time and group, F(6) = 1.34, p = .219, η² = .004. Thus, there were no indications of significant changes in subjective stress perception over time, between groups, or in their interaction (see Table 4).

**Table 4.**
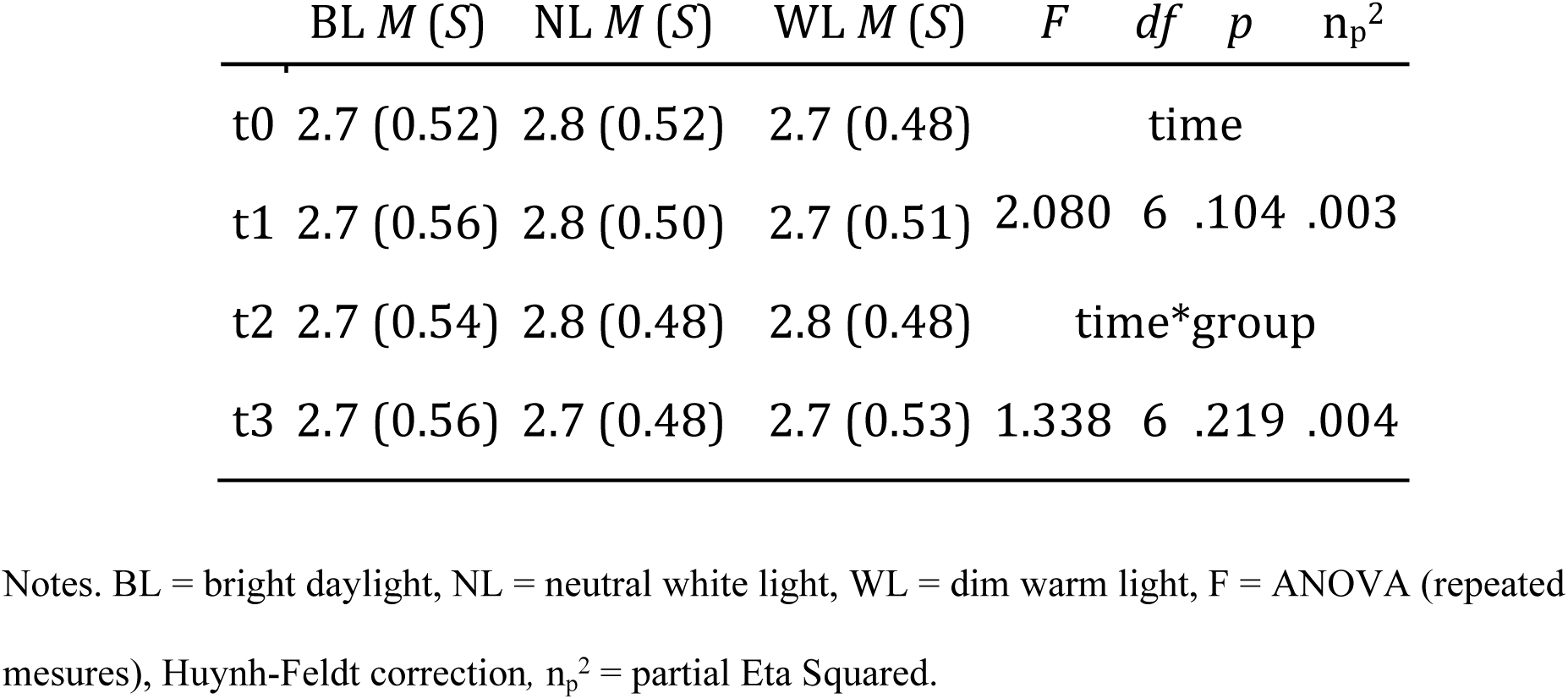
Analysis of perceived stress (n = 786).

## Discussion

### 4.1 Main Results

The results of the present study demonstrate significant effects of lighting conditions on two of the three attention components measured by the ANT (alerting and orienting), while only a non-significant trend was observed for executive attention. Although the BL lighting program appears to be generally advantageous, its difference from the NL condition is mostly small. This may be explained by the relatively low contrast between the two lighting programs in terms of illuminance and their very similar correlated color temperatures.

### 4.2 Limitations

The luminaires used in this study differed not only in their correlated color temperature and illuminance but also in other lighting-related parameters such as beam angle and brightness distribution, making it difficult to isolate the effects of individual variables. In contrast to Barkmann et al. [24], Auras et al. [23], and Slegers et al. [37], who each used a single light source, various luminaires were employed in the present study. Consequently, differences extended beyond illuminance and color temperature to include other photometric characteristics. Although this approach is not optimal from an experimental perspective, the study was conducted entirely without financial support from the lighting industry and was funded solely through private means by the authors. The use of different luminaires also reflects common practice in real-world application settings [3].

The contrast between the experimental lighting conditions may have been too small to detect clear effects. For future studies, a professionally controllable human-centric lighting (HCL) system has already been acquired to ensure more standardized and comparable lighting conditions.

Light measurements at the beginning of data collection were conducted in accordance with the standards in place at the time, using horizontal lux measurements, but without high-resolution spectrophotometric validation as is now recommended. Retrospective measurements were not possible because the test setup had already been dismantled at the time of statistical analysis.

The sample consisted primarily of university students who are accustomed to extended periods of concentration. Therefore, generalizability of the results to other populations is limited.

The ANT is considered robust against external influences such as ambient light [3]. However, in the present study, a more sensitive testing instrument might have revealed stronger effects. Still, the ANT remains a well-validated tool that enables conclusions regarding the activation of specific neural networks and is generally appropriate for the research question at hand.

In the warm white lighting condition, the computer monitor acted as an additional light source with a neutral white background, thereby reducing the contrast to the daylight white condition. It is likely that this systematically led to an underestimation of the observed effects.

Although the test booths were largely shielded from external light, it is assumed that participants were exposed to natural daylight before testing. This may have resulted in unintended pre-activation, especially in the control condition. Kent et al. [53], for example, demonstrated a significant correlation between daylight exposure (measured via NASA satellite data) and cognitive performance. While no association with local weather data was found in the present study (not shown), it remains plausible that prior daylight exposure influenced the results.

### 4.3 The relationship of the present results with previous studies

The results of this study partially confirm previous findings but deviate from theoretical expectations in some aspects. They indicate that lighting has differential effects on distinct components of attention.

#### Executive attention

The poorest performance in executive attention was observed under warm white light (WL). Neutral light (NL) showed a slight, though non-significant, advantage over bright daylight white light (BL). This trend suggests that cooler light may support cognitive control processes - an interpretation consistent with studies indicating an activating effect of higher blue-enriched light on prefrontal networks. Compared to the pilot study by Wessolowski and Rahim [3], which reported a significant advantage of neutral light over warm white light (in a smaller sample), this effect could not be replicated in the present study. Interestingly, and contrary to theoretical expectations, NL tended to outperform BL. This contradicts the assumption of a linear relationship between color temperature (or blue content) and executive performance and instead points to a more complex mechanism, possibly moderated by stress levels or contextual factors.

#### Alerting component

A significant medium-sized effect was found in favor of bright daylight white light (BL) for the alerting component. This finding aligns with meta-analytic evidence by Mu et al. [46] suggesting an activating effect of blue-enriched light on the central nervous system and general vigilance. In contrast to the pilot study by Wessolowski and Rahim [3], which found no significant effect, the current sample showed a clearer response.

#### Orienting component

Surprisingly, warm white light (WL) showed a significant advantage in the orienting component, which assesses selective shifts of attention. Theoretically, a benefit from BL would have been expected - either due to enhanced cognitive activation or, assuming a stress-reducing effect at lower illuminance, possibly also under WL. However, consistently low stress scores and the absence of significant differences in subjective stress perception between conditions (see ancillary results) argue against this interpretation. Similarly, the pilot study by Wessolowski and Rahim [3] showed only a small, non-significant, theoretically expected advantage for BL.

The unexpected pattern suggests that warmer light - despite its typically calming connotation - may enhance certain aspects of selective attentional focus. It is conceivable that warm white light, by lowering arousal, facilitates more precise attentional targeting while supporting stimulus selection.

The observations suggesting that daylight interventions subjectively lead to increased **stress levels** could not be confirmed in this study, nor in the pilot study [3]. No effect on stress load was found. Presumably, the effect of light on the perception of stress depends on the level of arousal.

Overall, the findings confirm that light does not affect all attentional dimensions equally. The partially contradictory results in relation to established light-effect theories highlight the need for more differentiated explanatory models. The theoretical framework proposed by Wessolowski [6] (Fig 1) offers a suitable perspective: It integrates biologically non-visual mechanisms (e.g., ipRGC-mediated activation), psychological processes (e.g., subjective experience, expectations), and contextual factors (e.g., spatial design, usage situation), and should be further developed into a structural equation model in a subsequent publication based on measurement data from multiple experiments.

It is also noteworthy that bright daylight white light (BL) yielded at least medium performance levels across all three ANT components and the highest scores in the alerting component. This is consistent with theoretical expectations, suggesting that bright, daylight-like lighting may exert a general activating effect through conditioned associations with activity and performance readiness. The relatively small difference between BL and NL may be attributed to the limited contrast between the lighting conditions—a methodological limitation that should be addressed in future studies.

These findings underscore the importance of targeted lighting design, particularly within the framework of human centric lighting (HCL). It appears advisable to align lighting not only with circadian rhythms but also with task-specific and emotional demands. In the future, adaptive systems that consider situational factors such as stress levels or mood - e.g., through emotion recognition - could enable more differentiated and user-optimized lighting control.

### 4.4 Summary

The present study investigated the effects of different lighting conditions on three attentional networks using the Attention Network Test (ANT). The findings demonstrate that light can modulate cognitive processes - but not uniformly; rather, its effects vary depending on the specific component of attention.

Bright daylight white light (BL) emerged as the most beneficial condition overall, particularly for the alerting component, whereas warm white light (WL) was repeatedly associated with unfavorable outcomes in both executive attention and alerting. However, an unexpected advantage of WL in the orienting component suggests a more complex underlying mechanism that cannot be fully explained by existing biological or psychological models.

The fact that effects were observed only in contrast-based ANT measures (i.e., difference scores) and not in the raw values of individual conditions indicates a specific influence on network-related cognitive processes. Future studies should examine lighting programs with greater contrast and expand the methodological design to include physiological measures (e.g., beam angle, luminance distribution, pupillometry, EEG) as well as more differentiated psychological assessments.

To better understand the interaction between biological, psychological, and contextual factors, the development and validation of integrative models of lighting effects - such as the HCL-effect model proposed by Wessolowski (2024) - is essential. Light is more than a visual stimulus; it influences attention and cognitive performance. This insight should be systematically considered in the design of future work, learning, and living environments.

## Data Availability

All relevant data are within the manuscript and its Supporting Information files.

## Acknowledgments

We would like to thank all participants in the study, as well as the students who served as test administrators. We are particularly grateful to Prof. Dr. Mike Wendt for his expertise in attention measurement and to Prof. Dr. Tilo Strobach for supporting the research project. We also thank the Medical School Hamburg, especially its managing director Illona Renken-Olthoff, for the continuous support of the project.

## References

1. Berson DM, Dunn FA, Takao M. Phototransduction by Retinal Ganglion Cells That Set the Circadian Clock. Science. 2002;295: 1070–1073. doi:10.1126/science.1067262

2. Wessolowski N, Barkmann C, Stuhrmann LY, Schulte-Markwort M. Wirkung von Lichttherapie auf den Nachtschlaf von Kindern mit Schlafproblemen. Zeitschrift für Kinder- und Jugendpsychiatrie und Psychotherapie. 2019 [cited 14 Jul 2025]. Available: https://econtent.hogrefe.com/doi/10.1024/1422-4917/a000683

3. Wessolowski N, Rahim RJ. Is executive attention affected by environmental lighting conditions? Steinborn MB, editor. PLoS ONE. 2025;20: e0305998. doi:10.1371/journal.pone.0305998

4. Fan J, McCandliss BD, Fossella J, Flombaum JI, Posner MI. The activation of attentional networks. NeuroImage. 2005;26: 471–479. doi:10.1016/j.neuroimage.2005.02.004

5. Pinel JPJ. Biopsychology. Pearson;

6. Wessolowski N. Licht, Farbe und Psyche: Theorie, Forschung und praktische Empfehlungen. 2nd ed. Hamburg: epubli; 2024.

7. Wessolowski N. Wirksamkeit von Dynamischem Licht im Schulunterricht. doctoralThesis, Staats- und Universitätsbibliothek Hamburg Carl von Ossietzky. 2014. Available: https://ediss.sub.uni-hamburg.de/handle/ediss/5418

8. Goldstein EB. Sensation and Perception. Belmont, Calif: Wadsworth Pub Co; 2010.

9. Brainard GC, Hanifin JP, Greeson JM, Byrne B, Glickman G, Gerner E, et al. Action Spectrum for Melatonin Regulation in Humans: Evidence for a Novel Circadian Photoreceptor. J Neurosci. 2001;21: 6405–6412. doi:10.1523/JNEUROSCI.21-16-06405.2001

10. Hattar S, Liao H-W, Takao M, Berson DM, Yau K-W. Melanopsin- Containing Retinal Ganglion Cells: Architecture, Projections, and Intrinsic Photosensitivity. Science. 2002;295: 1065–1070. doi:10.1126/science.1069609

11. Knez I. Effects of indoor lighting on mood and cognition. Journal of Environmental Psychology. 1995;15: 39–51. doi:10.1016/0272-4944(95)90013-6

12. Knez I, Enmarker I. Effects of Office Lighting on Mood and Cognitive Performance And A Gender Effect in Work-xRelated Judgment. Environment and Behavior. 1998;30: 553–567. doi:10.1177/001391659803000408

13. Vandewalle G, Maquet P, Dijk D-J. Light as a modulator of cognitive brain function. Trends in Cognitive Sciences. 2009;13: 429–438. doi:10.1016/j.tics.2009.07.004

14. Sahin L, Wood BM, Plitnick B, Figueiro MG. Daytime light exposure: Effects on biomarkers, measures of alertness, and performance. Behavioural Brain Research. 2014;274: 176–185. doi:10.1016/j.bbr.2014.08.017

15. Okamoto Y, Rea MS, Figueiro MG. Temporal dynamics of EEG activity during short- and long-wavelength light exposures in the early morning. BMC Research Notes. 2014;7: 113. doi:10.1186/1756-0500-7-113

16. Baron RA, Rea MS, Daniels SG. Effects of indoor lighting (illuminance and spectral distribution) on the performance of cognitive tasks and interpersonal behaviors: The potential mediating role of positive affect. Motiv Emot. 1992;16: 1–33. doi:10.1007/BF00996485

17. Rea M. Light-Much More Than Vision. 2003. Available: https://www.semanticscholar.org/paper/Light-Much-More-Than-Vision-Rea/db8f5ba0be450279bce83054781b513a70fc35ff?p2df

18. Biner PM, Butler DL, Fischer AR, Westergren AJ. An Arousal Optimization Model of Lighting Level Preferences: An Interaction of Social Situation and Task Demands. Environment and Behavior. 1989;21: 3–16. doi:10.1177/0013916589211001

19. Cajochen C. Alerting effects of light. Sleep Medicine Reviews. 2007;11: 453–464. doi:10.1016/j.smrv.2007.07.009

20. Smolders KCHJ, de Kort YAW. Bright light and mental fatigue: Effects on alertness, vitality, performance and physiological arousal. Journal of Environmental Psychology. 2014;39: 77–91. doi:10.1016/j.jenvp.2013.12.010

21. Al-Karawi D, Jubair L. Bright light therapy for nonseasonal depression: Meta-analysis of clinical trials. J Affect Disord. 2016;198: 64–71. doi:10.1016/j.jad.2016.03.016

22. Askaripoor T, Motamedzade M, Golmohammadi R, Farhadian M, Babamiri M, Samavati M. Non-Image Forming Effects of Light on Brainwaves, Autonomic Nervous Activity, Fatigue, and Performance. Journal of Circadian Rhythms. 2018;16. doi:10.5334/jcr.167

23. Auras MI, Barkmann C, Niemeyer M, Schulte-Markwort M, Wessolowski N. Wirksamkeit von variablem Licht in der Kinder- und Jugendpsychiatrie. Zeitschrift für Kinder- und Jugendpsychiatrie und Psychotherapie. 2016 [cited 14 Jul 2025]. Available: https://econtent.hogrefe.com/doi/10.1024/1422-4917/a000409

24. Barkmann C, Wessolowski N, Schulte-Markwort M. Applicability and efficacy of variable light in schools. Physiology & Behavior. 2012;105: 621– 627. doi:10.1016/j.physbeh.2011.09.020

25. Blume C, Garbazza C, Spitschan M. Effects of light on human circadian rhythms, sleep and mood. Somnologie. 2019;23: 147–156. doi:10.1007/s11818-019-00215-x

26. Boubekri M, Cheung IN, Reid KJ, Wang C-H, Zee PC. Impact of Windows and Daylight Exposure on Overall Health and Sleep Quality of Office Workers: A Case-Control Pilot Study. J Clin Sleep Med. 2014;10: 603–611. doi:10.5664/jcsm.3780

27. Cajochen C. Alerting effects of light. Sleep Medicine Reviews. 2007;11: 453–464. doi:10.1016/j.smrv.2007.07.009

28. Chang A-M, Scheer FAJL, Czeisler CA. The human circadian system adapts to prior photic history. J Physiol. 2011;589: 1095–1102. doi:10.1113/jphysiol.2010.201194

29. Fleischer SE. Die psychologische Wirkung veränderlicher Kunstlichtsituationen auf den Menschen. Doctoral Thesis, ETH Zurich. 2001. doi:10.3929/ethz-a-004122647

30. Hébert M, Martin SK, Lee C, Eastman CI. The effects of prior light history on the suppression of melatonin by light in humans. J Pineal Res. 2002;33: 198–203. doi:10.1034/j.1600-079x.2002.01885.x

31. Kompier ME, Smolders KCHJ, Kort YAW de. Abrupt light transitions in illuminance and correlated colour temperature result in different temporal dynamics and interindividual variability for sensation, comfort and alertness. PLOS ONE. 2021;16: e0243259. doi:10.1371/journal.pone.0243259

32. Meyer PT, Specht MB, Wessolowski N. Schlafbezogene Metakognitionen im transdiagnostischen Kontext. Somnologie. 2024 [cited 14 Jul 2025]. doi:10.1007/s11818-024-00478-z

33. Pachito DV, Eckeli AL, Desouky AS, Corbett MA, Partonen T, Rajaratnam SM, et al. Workplace lighting for improving alertness and mood in daytime workers. Cochrane Database Syst Rev. 2018;3: CD012243. doi:10.1002/14651858.CD012243.pub2

34. Porcheret K, Wald L, Fritschi L, Gerkema M, Gordijn M, Merrrow M, et al. Chronotype and environmental light exposure in a student population. Chronobiol Int. 2018;35: 1365–1374. doi:10.1080/07420528.2018.1482556

35. Rosenthal NE, Sack DA, Carpenter CJ, Parry BL, Mendelson WB, Wehr TA. Antidepressant effects of light in seasonal affective disorder. Am J Psychiatry. 1985;142: 163–170. doi:10.1176/ajp.142.2.163

36. Rutten S, Vriend C, van den Heuvel OA, Smit JH, Berendse HW, van der Werf YD. Bright Light Therapy in Parkinson′s Disease: An Overview of the Background and Evidence. Parkinson’s Disease. 2012;2012: 767105. doi:10.1155/2012/767105

37. Sleegers P, Moolenaar N, Galetzka M, Pruyn A, Sarroukh B, van der Zande B. Lighting affects students’ concentration positively: Findings from three Dutch studies. Lighting Research & Technology. 2013;45: 159–175. doi:10.1177/1477153512446099

38. Smith KA, Schoen MW, Czeisler CA. Adaptation of human pineal melatonin suppression by recent photic history. J Clin Endocrinol Metab. 2004;89: 3610–3614. doi:10.1210/jc.2003-032100

39. Terman M, Terman JS. Light therapy for seasonal and nonseasonal depression: efficacy, protocol, safety, and side effects. CNS Spectr. 2005;10: 647–663; quiz 672. doi:10.1017/s1092852900019611

40. Wehr TA. The durations of human melatonin secretion and sleep respond to changes in daylength (photoperiod). J Clin Endocrinol Metab. 1991;73: 1276–1280. doi:10.1210/jcem-73-6-1276

41. Werth L, Steidle A, Hubschneider C, de Boer J, Sedlbauer K. Psychologische Befunde zu Licht und seiner Wirkung auf den Menschen – ein Überblick. Bauphysik. 2013;35: 193–204. doi:10.1002/bapi.201310058

42. Zhou T, Dang W, Ma Y, Hu C, Wang N, Zhang G, et al. Clinical efficacy, onset time and safety of bright light therapy in acute bipolar depression as an adjunctive therapy: A randomized controlled trial. Journal of Affective Disorders. 2018;227: 90–96. doi:10.1016/j.jad.2017.09.038

43. Effects of Office Lighting on Mood and Cognitive Performance And A Gender Effect in Work-xRelated Judgment - Igor Knez, Ingela Enmarker, 1998. [cited 14 Jul 2025]. Available: https://journals.sagepub.com/doi/10.1177/001391659803000408

44. Phototransduction by Retinal Ganglion Cells That Set the Circadian Clock | Science. [cited 16 Jul 2025]. Available: https://www.science.org/doi/10.1126/science.1067262

45. Golmohammadi R, Yousefi H, Safarpour Khotbesara N, Nasrolahi A, Kurd N. Effects of Light on Attention and Reaction Time: A Systematic Review. J Res Health Sci. 2021;21: e00529. doi:10.34172/jrhs.2021.66

46. Mu Y-M, Huang X-D, Zhu S, Hu Z-F, So K-F, Ren C-R, et al. Alerting effects of light in healthy individuals: a systematic review and meta-analysis. Neural Regeneration Research. 2022;17: 1929. doi:10.4103/1673-5374.335141

47. Weitbrecht WU, Bärwolff H, Lischke A, Jünger S. Wirkung der Farbtemperatur des Lichts auf Konzentration und Kreativität. Fortschr Neurol Psychiatr. 2015;83: 344–348. doi:10.1055/s-0035-1553051

48. Brickenkamp R. The d2 Test of Attention: Manual. Cambridge, Mass.: Hogrefe Publishing; 1998.

49. Faul F, Erdfelder E, Buchner A, Lang A-G. Statistical power analyses using G*Power 3.1: Tests for correlation and regression analyses. Behavior Research Methods. 2009;41: 1149–1160. doi:10.3758/brm.41.4.1149

50. Steyer R, Schwenkmetzger P, Notz P, Eid M. Entwicklung des Mehrdimensionalen Befindlichkeitsfragebogens (MDBF). Primärdatensatz. 2004 [cited 21 Jul 2025]. Available: https://rdc-psychology.org/de/steyer_2004-2

51. Wetter Rückblick Hamburg - Wetterdaten weltweit. In: WetterOnline [Internet]. [cited 22 Jul 2025]. Available: https://www.wetteronline.de/wetterdaten/hamburg?pcid=pc_rueckblick_data&gid=10147&pid=p_rueckblick_diagram&sid=StationHistory&iid=10147&month=11&year=2017&period=4&metparaid=SDLD

52. Wessolowski N. Einführung in die Forschungsmethodik: Wissenschaftliches Arbeiten und Statistik: mit praktischer Durchführung, Prüfungsfragen, Umgang mit Künstlicher Intelligenz & vielem mehr. 1st ed. Berlin: epubli; 2023.

53. Kent ST, McClure LA, Crosson WL, Arnett DK, Wadley VG, Sathiakumar N. Effect of sunlight exposure on cognitive function among depressed and non-depressed participants: a REGARDS cross-sectional study. Environmental Health. 2009;8: 34. doi:10.1186/1476-069X-8-34

54. Smolders KCHJ, de Kort YAW. Bright light and mental fatigue: Effects on alertness, vitality, performance and physiological arousal. Journal of Environmental Psychology. 2014;39: 77–91. doi:10.1016/j.jenvp.2013.12.010

